# Inflammasome activation in COVID-19 patients

**DOI:** 10.1101/2020.08.05.20168872

**Authors:** Tamara S. Rodrigues, Keyla S.G. de Sá, Adriene Y. Ishimoto, Amanda Becerra, Samuel Oliveira, Leticia Almeida, Augusto V. Gonçalves, Debora B. Perucello, Warrison A. Andrade, Ricardo Castro, Flavio P. Veras, Juliana E. Toller-Kawahisa, Daniele C. Nascimento, Mikhael H.F. de Lima, Camila M. S. Silva, Diego B. Caetite, Ronaldo B. Martins, Italo A. Castro, Marjorie C. Pontelli, Fabio C. de Barros, Natália B. do Amaral, Marcela C. Giannini, Letícia P. Bonjorno, Maria Isabel F. Lopes, Maíra N. Benatti, Rodrigo C. Santana, Fernando C. Vilar, Maria Auxiliadora-Martins, Rodrigo Luppino-Assad, Sergio C.L. de Almeida, Fabiola R. de Oliveira, Sabrina S. Batah, Li Siyuan, Maira N. Benatti, Thiago M. Cunha, José C. Alves-Filho, Fernando Q. Cunha, Larissa D. Cunha, Fabiani G. Frantz, Tiana Kohlsdorf, Alexandre T. Fabro, Eurico Arruda, Renê D.R. de Oliveira, Paulo Louzada-Junior, Dario S. Zamboni

**Author notes:** Correspondence: Dario S. Zamboni, Ph.D., Departamento de Biologia Celular e Molecular e Bioagentes Patogênicos. Av., Bandeirantes 3900, Ribeirão Preto, SP 14049-900 Brazil.

## Abstract

Severe cases of COVID-19 are characterized by a strong inflammatory process that may ultimately lead to organ failure and patient death. The NLRP3 inflammasome is a molecular platform that promotes inflammation via cleavage and activation of key inflammatory molecules including active caspase-1 (Casp1p20), IL-1β and IL-18. Although the participation of the inflammasome in COVID-19 has been highly speculated, the inflammasome activation and participation in the outcome of the disease is unknown. Here we demonstrate that the NLRP3 inflammasome is activated in response to SARS-CoV-2 infection and it is active in COVID-19, influencing the clinical outcome of the disease. Studying moderate and severe COVID-19 patients, we found active NLRP3 inflammasome in PBMCs and tissues of post-mortem patients upon autopsy. Inflammasome-derived products such as Casp1p20 and IL-18 in the sera correlated with the markers of COVID-19 severity, including IL-6 and LDH. Moreover, higher levels of IL-18 and Casp1p20 are associated with disease severity and poor clinical outcome. Our results suggest that the inflammasome is key in the pathophysiology of the disease, indicating this platform as a marker of disease severity and a potential therapeutic target for COVID-19.

---

COVID-19 is an inflammatory disease caused by the severe acute respiratory syndrome coronavirus 2 (SARS-CoV-2), which can manifest a broad spectrum of symptoms ranging from little or no symptoms to severe pneumonia that may evolve to acute respiratory distress syndrome (ARDS) and death^1^. While the molecular mechanisms driving disease severity are still unclear, the clinical association of inflammatory mediators such as IL-6 and lactate dehydrogenase (LDH) with severe cases suggests that excessive inflammation is central for the poor clinical outcome ^2-5^. The induction of inflammatory processes in the host cell often requires the engagement of the inflammasomes, which are protein platforms that aggregate in the cytosol in response to different stimuli ^6^. The NLRP3 inflammasome, possibly the most studied one of such platforms, comprises the NLRP3 receptor, the adaptor molecule ASC, and caspase-1. Caspase-1 is activated by proteolytically cleavage and promotes the activation of many substrates, including the inflammatory cytokines IL-1β and IL-18 and Gasdermin-D, a pore-forming protein that induces an inflammatory form of cell death called pyroptosis ^6^. NLRP3 activation in response to microbial infections, cell damage, or aggregates in the host cell cytoplasm promotes ASC polymerization, leading to the formation of a micron-sized structure called puncta (or speck) that is a hallmark of active inflammasomes in the cells ^7^.

The pronounced inflammatory characteristics of COVID-19 and the correlation of disease severity with the pyroptosis marker LDH prompted us to investigate the activation of the inflammasome by SARS-CoV-2 and its role in disease development. Initially, we infected primary human monocytes in vitro with SARS-CoV-2 and assessed inflammasome activation. Using monocytes from different healthy donors, we found that SARS-CoV-2 infection triggers LDH release and activation of NLRP3 inflammasome in monocytes, as shown by NLRP3 puncta formation (**Fig. 1A-C**). As expected, uninfected cells were negative for dsRNA staining and NLRP3 puncta formation. Nigericin, a bacterial toxin known to trigger NLRP3 inflammasome, was used as a positive control. Activation of the NLRP3 inflammasome required viable virus particles, as UV-inactivated SARS-CoV-2 failed to induce NLRP3 puncta formation (**Fig. 1B**). We also measured the activation of IL-1β as a readout for inflammasome activation and found that infection with SARS-CoV-2 caused the production of IL-1β in primed cells, as measured by ELISA (**Fig. 1D**). We measured the viral load using RT-PCR and confirmed that SARS-CoV-2 infects and replicates in primary human monocytes in vitro (**Fig. 1E**). We also tested infection in primary human monocyte-derived macrophages and found that SARS-CoV-2 infection triggers macrophage cell death in a dose-dependent effect as shown by the presence of LDH in the supernatants (**Fig. 1F**). Nigericin was used as positive control and UV-inactivated virus as a negative control. We next tested the inflammasome activation in the sera of COVID-19 patients. It was previously shown that IL-1β is activated independently of caspase-1 in vivo ^8-10^, thus we assessed active/cleaved caspase-1 (Casp1p20) and cleaved IL-18 as readouts for inflammasome activation in COVID-19 patients. We tested sera from 124 COVID-19 patients obtained on the day of hospitalization (all tested RT-PCR positive for SARS-CoV-2) and compared with sera of 42 controls that tested RT-PCR/serology negative or that were collected before the COVID-19 pandemic. We found higher concentrations of Casp1p20 and IL-18 in the sera of patients (Fig. 2A, B), suggesting active inflammasome in COVID-19 patients. We also found that IL-6, IL-10, IL-4 were increased in patients as compared to controls, whereas we do not detect statistically significant differences for IL-2, TNF-α, and IL-17A (**Fig. 2C and Fig. S1**). IFN-γ levels were slightly lower in COVID-19 patients as compared to controls (**Fig. S1**). Next, we measured inflammasome activation in peripheral blood mononuclear monocytes (PBMCs) from 47 patients and compared them with 32 healthy individuals. Using the FAM-YVAD assay that stains active intracellular caspase-1^11^, we found that, on the day of hospitalization, the PBMCs from patients show a higher percentage of FAM-YVAD+ cells as compared to healthy controls (**Fig. 2D-E**). Microscopy observation of these cells allows clear visualization of NLRP3 puncta in PBMCs, indicating active inflammasomes in patients cells (**Fig. 2F-G**). We further confirmed caspase-1 activation using a luminescent assay and found active caspase-1 in supernatants from 46 patients PBMCs cultures, as opposed to low caspase-1 activation in healthy donors (Fig. 2H). We also detected IL-1β in the supernatants of PBMCs from some patients, but not from healthy donors (**Fig. 2I**), further supporting inflammasome activation in PBMCs from COVID-19 patients.

**Figure 1.**
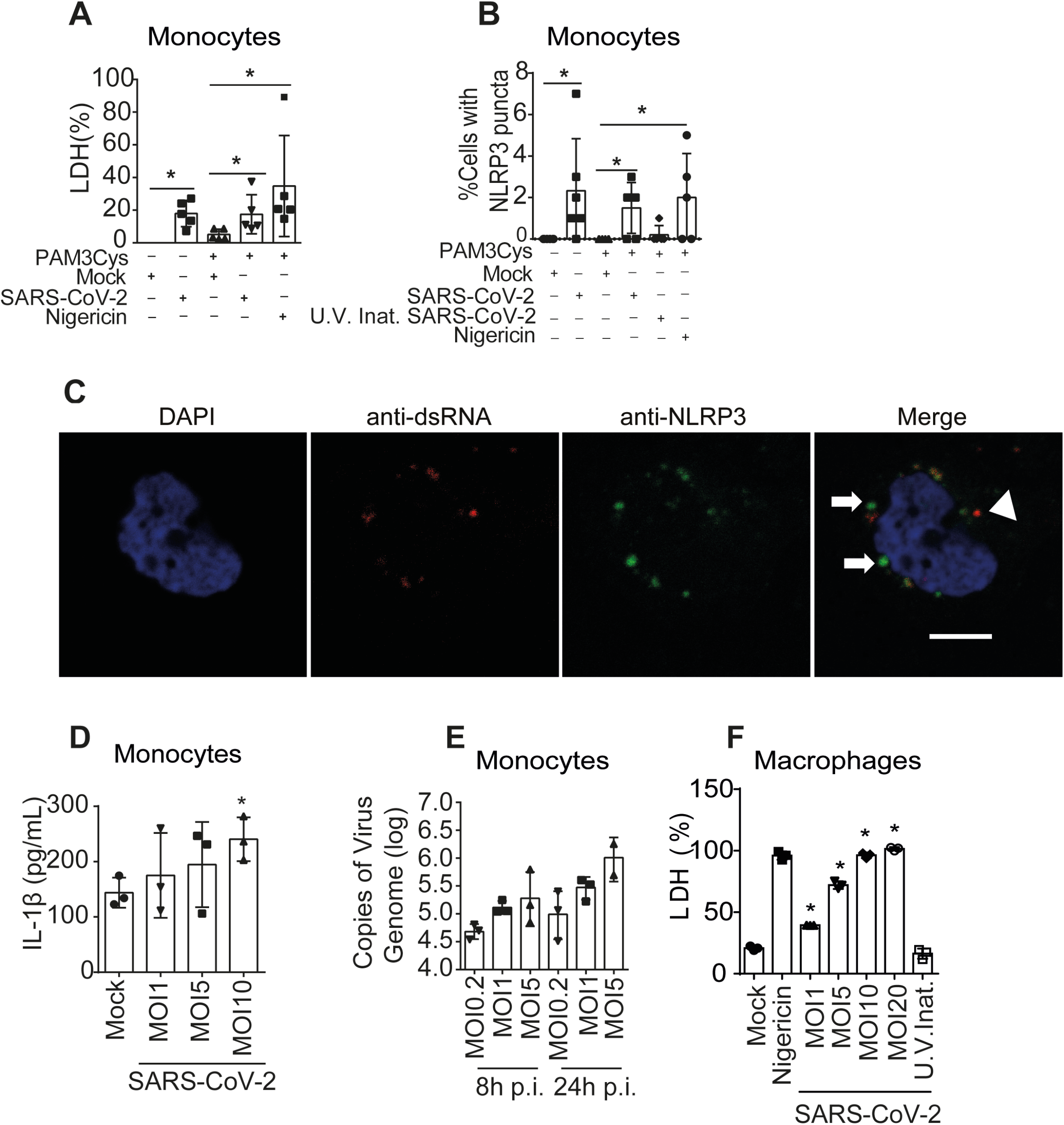
Infection of primary human monocytes with SARS-CoV-2 triggers inflammasome activation. **(A-E)** Human CD14+ monocytes were primed or not with PAM3Cys (300ng/mL) for 4 hours and infected with SARS-CoV-2 at a multiplicity of infection (MOI) of 0.2, 1 an 5 for 24 h. Mock was used as a negative infection control, and nigericin as a positive NLRP3 activation control. (A) LDH release was measured in the supernatants from 5 different donors. Triton (9%) was used to induce cell death and estimate 100% death. (B) the percentage of cell containing NLRP3 puncta was estimated in cells from 5 different donors. (C) A representative image of a monocyte containing NLRP3 puncta (green, indicated by arrows) and replicating SARS-CoV-2, depicted by anti-dsRNA antibody staining (red, indicated by an arrowhead) is shown. Nuclei stained in blue. Scale bar 5 μm. (D) IL-1β production was analyzed in the tissue culture supernatants of monocytes infected or not infected (MOCK) with the indicated MOI in experimental replicates. (D) Viral loads in the cell culture supernatants were estimated by RT-PCR in monocytes infected for 8 and 24 hs at the indicated MOI. (F) Monocytes from one donor were derived into macrophage and primed with PAM3Cys (300ng/mL) for 4 hours and infected with SARS-CoV-2 at a MOI of 1, 5 and 10 and 20 for 24 h. LDH were measured in the supernatants of experimental replicates. Mock and UV-irradiated virus (U.V. Inat.) was used as a negative infection control, and nigericin as a positive control. * *P* < 0.05, as determined by Student’s t-test. Box shows the average ± SD of the values.

**Figure 2.**
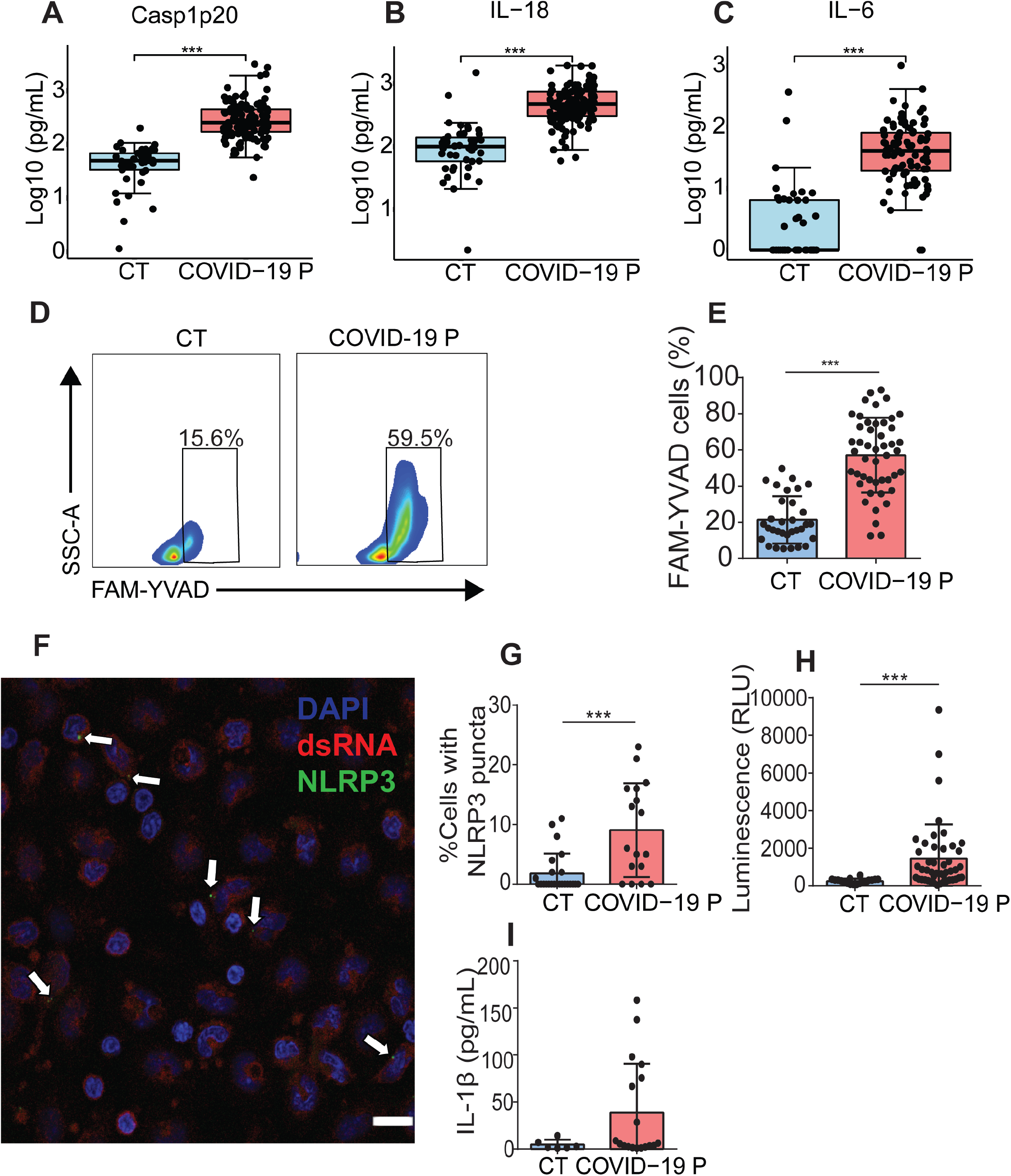
Inflammasome activation in COVID-19 patients. (**A-C**) Cytokine concentration in the serum control individuals (CT, n=42 to ELISA and 45 to CBA) and COVID-19 patients (COVID-19 P, n=124 to ELISA and 92 to CBA; all tested positive using RT-PCR). Active caspase-1 (Casp1p20, A) and IL-18 (B) were measured by ELISA, and IL-6 (C) were measured by CBA. Data are shown as Log10-transformed concentrations in pg/mL. (D-I) Peripheral Blood Mononuclear Cells (PBMCs) were isolated from fresh blood of CT or COVID-19 P. (D-E) FAM-YVAD positive PBMCs were estimated by FACS using FLICA Caspase-1 Assay Kit. (D) Representative histograms or one representative CT and one COVID-19 P indicate the gate for determination of the percentage of FAM-YVAD+ cells. (E) The percentage of FAM-YVAD+ cells for the 32 CT and 47 COVID-19 P. (F) PBMCs from COVID-19 P were stained with anti-dsRNA (red, indicating SARS-CoV-2 replication) and anti-NLRP3 (green) for determination of NLRP3 puncta (indicated by white arrows). Dapi (blue) stains the nuclei. Scale bar 20 μm. (G) The percentage of cells with NLRP3 puncta are shown for 24 CT and 17 COVID-19 P. (H) PBMCs from 18 CT and 46 COVID-19 P were maintained in culture for 16 hours and the supernatants were assayed for caspase-1 activity using the Caspase-Glo 1 Assay (H). (I) PBMCs from 6 CT or 18 COVID-19 P were maintained in culture for 16 hours and IL-1β production were estimated by ELISA. * *P* < 0.05, ** *P* < 0.01 and *** *P* < 0.001 as determined by Student’s t-test or Mann Whitney. Each dot represents the value form a single individual. Box shows average ± SD of the values.

We further assessed inflammasome activation in lung tissues obtained from autopsies of deceased COVID-19 patients. Using an anti-SARS-CoV-2 antibody, we first confirmed viral presence by immunohistochemical staining SARS-CoV-2 in injured regions of post-mortem lung tissues (**Fig. 3A,B**). We then performed Multiplex Staining by Sequential Immunohistochemistry with SARS-CoV-2, anti-CD14, and anti-NLRP3 and identified infected CD14+ cells expressing NLRP3 in post-mortem tissues (**Fig. 3C, D**). Using multiphoton microscopy, we quantified the number of NLRP3 puncta in tissues 5 controls and 6 COVID-19 patients and found that patients contain higher numbers of NLRP3 puncta as compared to controls. (**Fig. 3E**). Images of tissues stained with anti-NLRP3 antibody illustrates NLRP3 puncta in of lethal cases of COVID-19 (**Fig. 3F-I**). We observed NLRP3 puncta inside cells in the tissues and also cells contained in venules (**Fig. 3I**), unequivocally demonstrating activation of the NLRP3 inflammasome in fatal cases of COVID-19.

**Figure 3.**
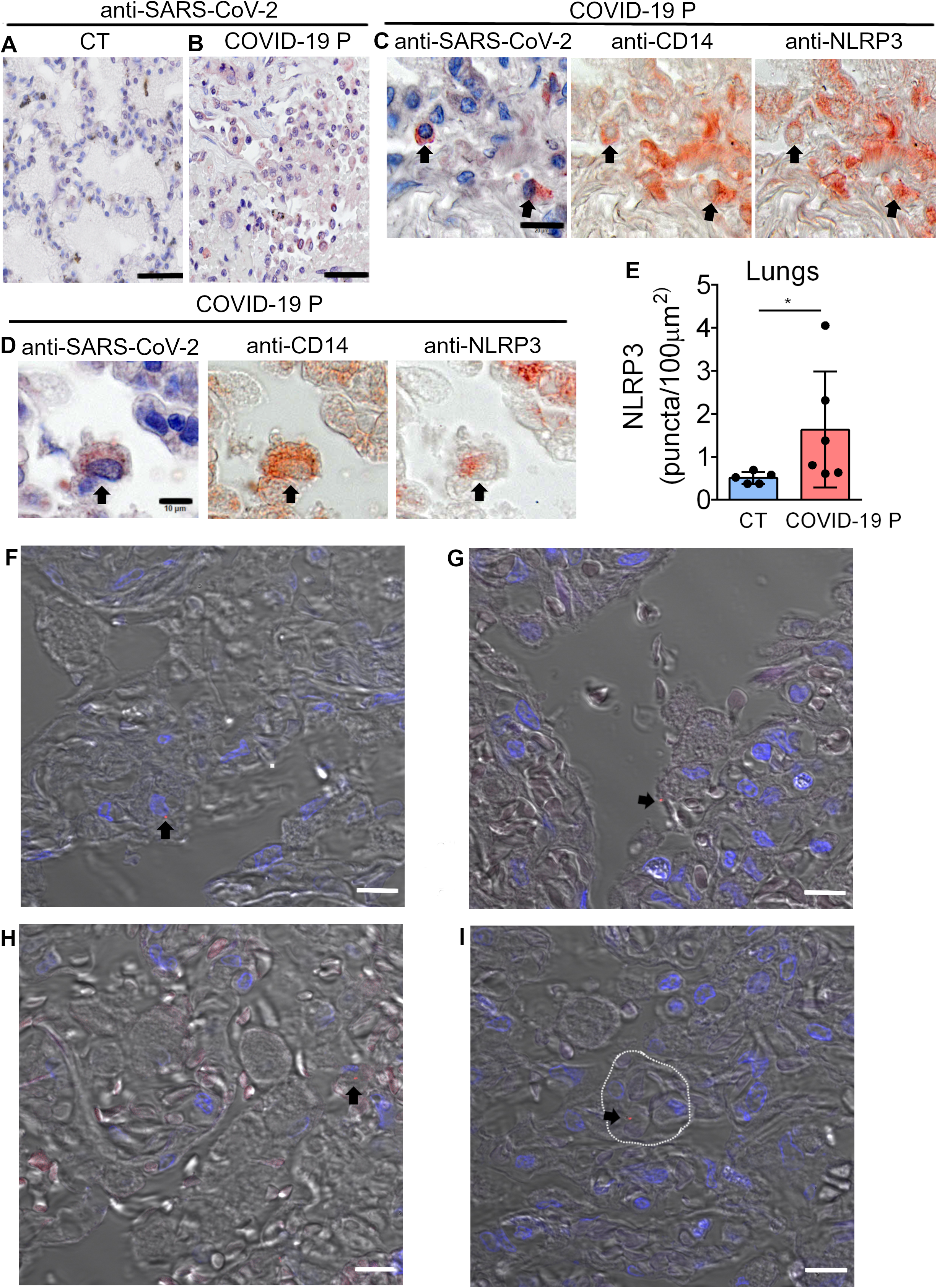
Lung histopathological analysis and NLRP3 activation in fatal cases of COVID-19. Representative pulmonary histological findings in COVID-19 patient (COVID-19 P), autopsied by ultrasound guided-minimally invasive autopsy. (A, B) Representative Immunohistochemical image of tissues from Control (CT, A) or COVID-19 P (B) stained with anti-SARS-CoV-2. Scale bar 50 μm. (C-D) Multiplex staining by sequential immunohistochemistry staining with anti-SARS-CoV-2, anti-CD14 and anti-NLRP3 arrows indicates infected CD14+ cells expressing NLRP3. Scale bar 20 μm (C) and 10 μm (D). (E) Quantification of NLRP3 puncta in pulmonary tissues of 5 CT and 6 COVID-19 P. (F, I) Multiphoton microscopy of tissues stained with anti-NLRP3 antibody indicates NLRP3 puncta (red, indicated by black arrows) in the pulmonary tissues. DAPI stains nuclei (blue). (I) NLRP3 puncta in a cell inside a venule (dotted white line). Scale bar 10 μm.

We next assessed the impact of the inflammasome activation in the clinical outcome of the disease. Initially, we performed analyses using the levels of Casp1p20 and IL-18 in 124 patients sera obtained on the day of hospitalization. A correlation matrix show association of Casp1p20 and IL-18 levels with patient characteristics and clinical parameters (**Fig. 4A**). As expected, we found a positive correlation between Casp1p20 and IL-18 levels (**Fig. 4B**). In addition, Casp1p20 positively correlated with IL-6, LDH and C-reactive protein (CRP) (**Fig. 4C-E**). Furthermore, we found that IL-18 levels positively correlated with IL-6 and CRP levels (**Fig. 4F, G**). We also evaluated if the levels of Casp1p20 and IL-18 were affected by patients comorbidities and clinical parameters, including bacterial co-infections (cultivable bacteria in the blood), nephropathy, obesity, gender, cerebrovascular accident, pneumopathy, immunodeficiency and neoplasia (**Fig. S2**). We only detected statistically significant differences when we compared obese with non-obese patients, as the levels of IL-18 were higher in patients with body mass index ≥ 30 (**Fig. S2G**).

**Figure 4.**
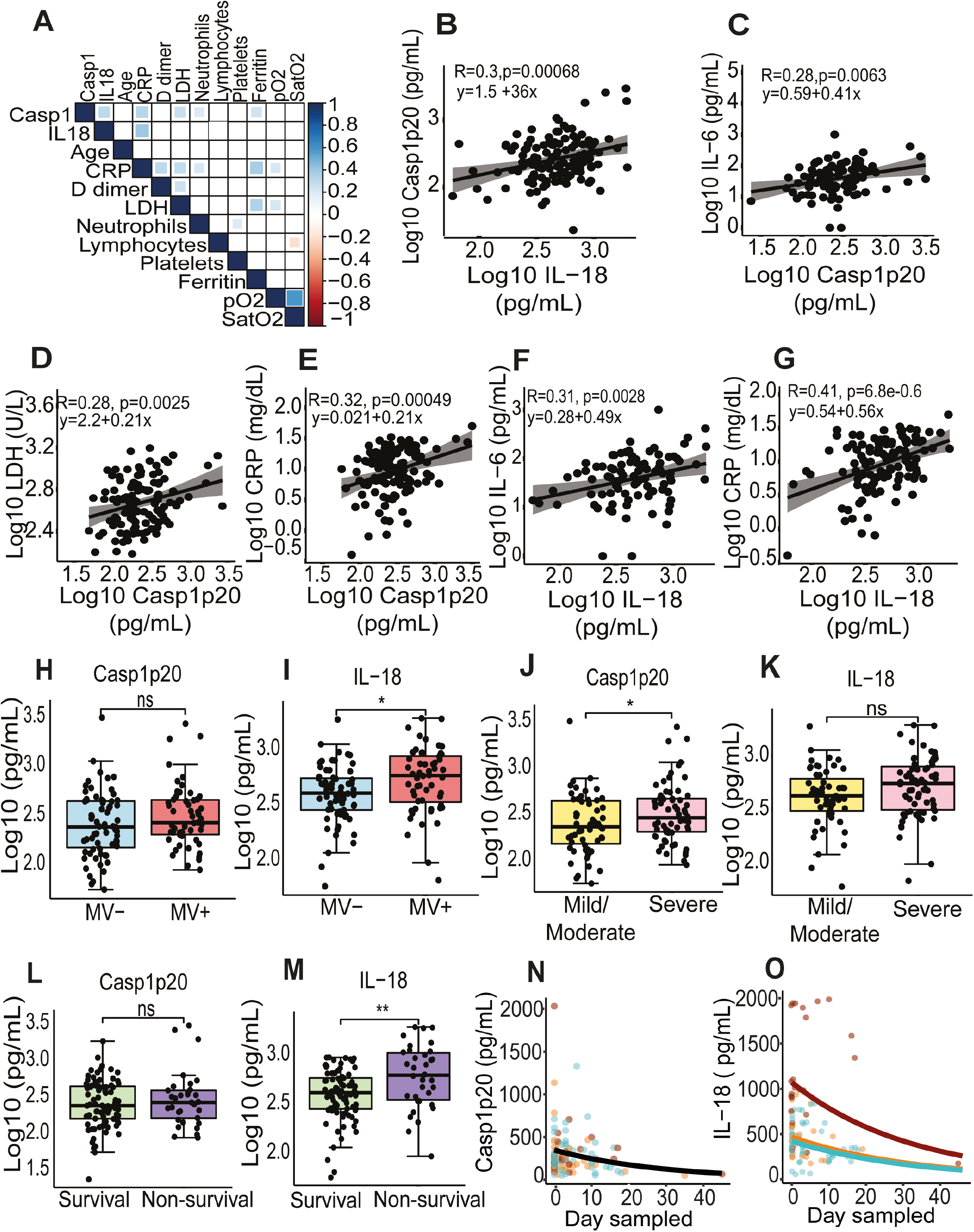
Inflammasome activation influences the clinical outcome of COVID-19. (A) Correlation matrix of Casp1p20 and IL-18 levels in the serum of COVID-19 patients at the hospitalization day with patient characteristics and clinical parameters. (B-J) Correlations of Casp1p20 with IL-18 (B), Casp1p20 with IL-6 (C), Casp1p20 with lactate dehydrogenase (LDH) (D), Casp1p20 with C-reactive protein (CRP), IL-18 with IL-6 (F) and IL-18 with CRP (G). (H,I) Levels of Casp1p20 (H) and IL-18 (I) in patients that required (MV+, blue box) or not (MV-, red box) mechanical ventilation. (J,K) Levels of Casp1p20 (J) and IL-18 (K) in patients with Mild/Moderate (yellow box) or Severe COVID-19 (pink box). (L,M) Levels of Casp1p20 (L) and IL-18 (M) in survivors (green box) or non-survivors (purple box). The levels of Casp1p20 and IL-18 were measured by ELISA and are shown as Log10-transformed concentrations in pg/mL. * *P* < 0.05, ** *P* < 0.01 and *** *P* < 0.001 as determined by Student’s t-test. Each dot represents value to form a single individual. Box shows average ± SD of the values. (N, O) Derived predictions from the best-fit models retained in Casp1p20 (N) and IL-18 (O) longitudinal analyses; IL-18 Model (O) comprises variation in the intercept among patients’ groups: Death (Red), Critical/Recovery (orange) and Mild/Recovery (blue).

Next, we investigated if Casp1p20 and IL-18 levels measured on the day of hospitalization correlated with the clinical outcome of the disease. Importantly, we found that IL-18, but not Casp1p20 were higher in patients who required mechanical ventilation (MV) as compared with patients that did not (**Fig. 4H, I**). When we separated the patients according to the severity of the disease (mild/moderate versus severe), we found that the levels of Casp1p20 but not IL-18 were higher in patients with the severe form of COVID-19 (**Fig. 4J, K**). We also observed that the levels of IL-18, but not Casp1p20, were higher in lethal cases of COVID-19 as compared to survivors (**Fig. 4L, M**). Finally, we performed longitudinal analyses of IL-18 and Casp1p20 production in 37 patients from the day of hospitalization (day zero) for up to 45 days post-admission using generalized mixed models with ‘gamma’ distribution and ‘log’ link function as the best dataset prediction. For these analyses, we categorized the patients as Death, Mild-Recovery (patients that were hospitalized but did not require mechanical ventilation and recovered), and Critical-Recovery (patients that required mechanical ventilation at the Intensive Care Unit (ICU) and recovered) (Supplemental data). For Casp1p20, the most parsimonious model indicated a significant effect of the day sampled, and the production level decreased with time regardless of sex or patient outcome (**Fig. 4N**, and **Supplemental data**). For IL-18, the best-fit model retained also described an overall reduction in IL-18 production along time (Day sampled), which differed among patient groups. This model predicted a decrease in IL-18 at similar rates among the three groups, with patients that died presenting higher levels (intercept) that never reached those observed in mild and critical recovered patients (**Fig. 4O** and **Supplemental data**), supporting our assertion that the magnitude of inflammasome activation impacts the disease outcome. In summary, our data demonstrate that the inflammasome is robustly active in COVID-19 patients requiring hospitalization. It also supports that both the magnitude of inflammasome activation at the hospitalization day and the course of inflammasome activation during hospitalization influenced the clinical outcome. Collectively, our observations suggest that the inflammasome is key for the induction of the massive inflammation observed in severe and fatal cases of COVID-19. Our study supports the use of inflammasome activation both as a marker of disease severity and prognostic but also as a potential therapeutic target for COVID-19.

## Materials and methods

### Patients

A total of 124 patients with COVID-19 that were tested positive using RT-PCR as described previously ^12,13^. Patients were classified according to their clinical manifestations in: i) mild cases: the clinical symptoms are mild and no pneumonia manifestations can be found in imaging; ii) moderate cases: patients have symptoms such as fever and respiratory tract symptoms, etc. and pneumonia manifestations can be seen in imaging; iii) severe cases: adults who meet any of the following criteria: respiratory rate ≥30 breaths/min; oxygen saturations; 93% at a rest state; arterial partial pressure of oxygen (PaO_2_)/oxygen concentration (FiO_2_) < 300 mm Hg^14^. Patients were enrolled in HC-FMRP/USP from April 06 to July 02, 2020 and **Table S1** summarizes clinical, laboratory, and treatment records. We also collected samples from 73 age and gender-matched healthy controls. Controls were collected either before the COVID-19 pandemic or tested negative for COVID-19 using RT-PCR and/or serology (specific IgM and IgG antibodies) (Asan Easy Test.COVID-19 IgM/IgG kits, Asan Pharmaceutical Co.).

### Peripheral blood mononuclear cells isolation

Whole blood was collected from healthy donors (Ethics Committee Protocol from the Clinical Hospital of Ribeirão Preto-USP: CAAE, n° 06825018.2.3001.5440) in tubes containing EDTA (BD Vacutainer CPTTM), according to the manufacturer’s instructions. The material was centrifuged at 400 x g for 10 minutes at room temperature. Then, the plasma was discarded and the cell pellet was resuspended in PBS 1X pH 7.4 (GIBCO, BRL). The cells were applied to the Ficoll-PaqueTM PLUS gradient column (GE Healthcare Biosciences AB, Uppsala, Sweden). Then, they were centrifuged at 640 x g for 30 minutes at room temperature to obtain the purified mononuclear fraction, which was carefully collected and transferred to a new tube. The cells were washed and the pellet was resuspended in RPMI for the subsequent analysis.

### Purification of monocytes from healthy donors and differentiation into macrophages

The PBMCs were quantified and the monocytes (CD14+ cells) were purified using positive selection with magnetic nanoparticles (BD). Briefly, PBMCs were labeled with BD IMag™ Anti-human CD14 Magnetic Particles - DM. The cells were transferred to a 48-well culture plate and placed over a magnetic field of the cell separation. Labeled cells migrated toward the magnet (positive fraction) whereas unlabeled cells were drawn off (negative fraction). The plate was then removed from the magnetic field for resuspension of the positive fraction. The separation was repeated twice to increase the purity of the positive fraction. The CD14 + monocytes resulting cells from this process were used for experiments or cultured in RPMI 1640 (GIBCO, BRL) containing 10% SFB and 50 ng/mL GM-CSF (R&D Systems) for 7 days for differentiation into macrophages.

### Virus stock production and in vitro infection

The SARS-CoV-2 used was Brazil/SPBR-02/2020 strain, isolated from the first Brazilian case of COVID-19. Viral stock was propagated under BSL3 conditions in Vero E6 cells, cultured in Dulbecco minimal essential medium (DMEM) supplemented with heat-inactivated fetal bovine serum (10%) and antibiotics/antimycotics (Penicillin 10,000 U/mL; Streptomycin 10,000 μg/mL). For preparation of viral stocks, Vero cells were infected in the presence of trypsin-TPCK (1μg/μL) for 48 hours at 37°C in a 5% CO_2_ atmosphere. When the virus-induced cytopathic effect, the cells were harvested with cell scrapers, and centrifuged (10.000 x G). The supernatant was stored at -80°C, and the virus titration was performed in Vero cells using standard limiting dilution to confirm the 50% tissue culture infectious dose (TCID50).

For human cells infections, 2×10^5^ purified human monocytes or monocyte derived macrophages were plated in 48 well plates, and infected with SARS-CoV-2 at Multiplicity of Infection (MOI) of MOI0.2, MOI1, and MOI5. After 2 hours of viral infection, the cells were washed with PBS1x, and a new medium (RPMI 10% FBS without Fenol Red) was added. Cells were incubated for 24h at 37°C in the presence of 5% CO_2_ atmosphere. After incubation, cells were processed for immunofluorescence assays and the supernatant was collected for determination of viral loads, cytokine production and LDH quantification.

### RT-PCR for SARS-CoV-2

Detection of SARS-CoV-2 was performed with primer-probe sets for 2019-nCoV_N2 and gene E, according to USA-CDC and Charité group protocols ^12,13^. The genes evaluated (N2, E, and RNAse-P housekeeping gene) were tested by one-step real-time RT-PCR using total nucleic acids extracted with Trizol® (Invitrogen, CA, EUA) from 50μL of cells supernatants in order to measure the genome viral load from the in vitro assays. All real-time PCR assays were done on Step-One Plus real-time PCR thermocycler (Applied Biosystems, Foster City, CA, USA). Briefly, RNA extraction was performed by Trizol®. A total of 100 ng of RNA was used for genome amplification, adding specifics primers (20 μM), and probe (5 μM), and with TaqPath 1-Step qRT-PCR Master Mix (Applied Biosystems, Foster City, CA, USA), with the following parameters: 25°C for 2 min, 50°C for 15 min, 95°C for 2 min, followed by 45 cycles of 94 °C for 5 s and 60 °C for 30s. Primers used were: N2 fwd: 5’-TTA CAA ACA TTG GCC GCA AA-3’; N2 rev: 5’-GCG CGA CAT TCC GAA GAA-3’; N2 probe: 5’-FAM-ACA ATT TGC CCC CAG CGC TTC AG-BHQ1-3’^13^; E fwd: 5’-ACA GGT ACG TTA ATA GTT AAT AGC GT-3’; E rev: 5’-ATA TTG CAG CAG TAC GCA CAC A-3’; E probe: 5’-AM-ACA CTA GCC ATC CTT ACT GCG CTT CG-BHQ-1-3’ ^12^; RNAse-P fwd: 5’-AGA TTT GGA CCT GCG AGC G-3’; RNAse-P rev: 5’-GAG CGG CTG TCT CCA CAA GT-3’; RNAse-P probe: 5’-FAM-TTC TGA CCT GAA GGC TCT GCG CG - BHQ-1-3’ ^13^.

### Evaluation of active caspase-1 activity and LDH release in cultured cells

For LDH determination, 2 × 10^5^ human CD14+ cells or human monocyte derived-macrophages were plated on 48-well plates in RPMI 10% FBS and incubated overnight. In the following day, cells were infected with SARS-CoV-2 using MOI 0.2, MOI 1, and MOI 5 in RPMI without Phenol Red (3.5 g/L HEPES, 2 g/L NaHCO_3_, 10.4 g/L RPMI without Phenol Red, 1% glutamine, pH 7.2) and incubated for 24 h. The supernatant was collected and LDH release was measured using CytoTox 96® Non-Radioactive Cytotoxicity Assay (Promega, Winsconsin, USA) following the manufacturer’s instructions. To evaluate caspase-1 activation, 5 × 10^5^ PBMC from COVID-19 patients or healthy donors were centrifuged (400g, 10 minutes) and cells were labeled for 30 minutes with the FLICA carboxyfluorescein reagent (FAM – YVAD – FMK, Immunochemistry Technologies, LLC), as recommended by the manufacturer. The cells were then washed two times with PBS 1x, and fixed with fixative reagent provided by manufacture. Acquisition was performed in fixed cells in flow cytometer (BD Accuri™ C6) and then analyzed using “FlowJo” (Tree Star, Ashland, OR, USA) software. To evaluate caspase-1 activity in supernatants, 2×10^5^ PBMCs were plated in 96 wells plate, and incubated overnight. To measure caspase-1 activity, the supernatants were collected, and incubated with the Luciferin WEHD-substrate provided by the Caspase-Glo 1 Assay (Promega). After 1 hour incubation at room temperature, luminescence was measured using SpectraMax i3 system (Molecular Devices).

### Immunofluorescence staining of isolated cells

For staining PBMCs from COVID-19 patients, a total of 5×10^5^ PBMCs were plated in 8 wells chamber slides for 1 h in RPMI without FBS for cell adhesion before fixation. For staining cells infected in vitro a total of 2×10^5^ human monocytes or monocyte differentiated macrophages were plated in 24-wells plate containing coverslips and infected with SARS-CoV-2 at indicated MOI for 16h. For fixation of the samples, tissue culture supernatants were removed and cells were fixed with 4% paraformaldehyde (PFA) for 20 minutes at room temperature. PFA was removed, cells were washed with PBS1x, and the coverslips or chambers were processed for IF as described. Cells were blocked and permeabilized using PBS 1x with goat serum and 0.05% saponin for 1h at room temperature. Primary antibodies mix of rabbit mAb anti-NLRP3 (Cell Signaling, 1:1000) were diluted in blocking solution and added to each chamber/coverslip. After 1h of incubation the samples were washed with PBS 1x and secondary antibodies were added and incubated for 1h at room temperature. Secondary antibodies used were goat anti-rabbit 488 (Invitrogen,1:3000) and goat anti-rabbit 594 (Life Technologies,1:3000). Slides were washed and mounted using DAPI (1mM) and ProLong (Invitrogen).

### Lung samples from autopsies and immunofluorescence and imaging

Adapted minimally invasive autopsies were performed in COVID-19 patients ^15-17^ Briefly, a mini thoracotomy (3cm) was done under the main area of lung injury identified by prior x-ray or computed tomography. The lung parenchyma is clamped by Collins Forceps, cut and fixed in 10% buffered formalin. Pulmonary tissue samples were stained with hematoxylin and eosin (H&E) and immunostaining as reported ^18 19^. The slides were incubated with the primary antibodies, rabbit anti-CD14 mAb (Abcam, 1:200) and mouse anti-Nlrp3 mAb (AdipoGen, 1:200), for 2h at room temperature or overnight at 4°C. Goat anti-mouse Alexa fluor-647 (Invitrogen) or goat anti-rabbit Alexa fluor-594 (Invitrogen) were used as secondary antibodies. Images were acquired by Axio Observer combined with LSM 780 confocal device with 630 x magnification (Carl Zeiss). Minimally invasive autopsies were approved by the FMRP/USP Ethical Committee (protocol #4.089.567).

### Sequential immunoperoxidase labeling and erasing

Tissue sections from paraffin-embedded lung fragments obtained from COVID-19 fatal cases were tested by immunohistochemistry (IHC) using anti-SARS-CoV-2 polyclonal antibody for in situ detection of SARS-CoV-2. Sequential immunoperoxidase labeling and erasing (SIMPLE) was then performed to determine additional markers after SARS-CoV-2 immune stain, using antibodies to CD14 (Abcam, 1:100 dilution), NLRP3 (Cell Signaling, 1:100 dilution) (Glass et al., 2009). After the incubation with primary antibody, the slides were incubated with immune-peroxidase polymer anti-mouse visualization system (SPD-125, Spring Bioscience, Biogen) and then with the chromogen substrate AEC peroxidase system kit (SK-4200, Vector Laboratories, Burlingame, CA). Microphotographs after immunostaining of tissue slides were scanned on a VS120 Olympus. After high-resolution scanning, slides coverslips were removed in PBS and dehydrated through ethanol gradient to 95% ethanol. Slides were incubated in ethanol series until erasing AEC color reaction. Following rehydration, antibodies were eluted by incubating sections in 0.15 mM KMnO_4_/0.01 M H_2_SO_4_ solution for 2 min, followed immediately by a distilled water wash. Tissue was then restained as indicated in the blocking step.

### Cytokine quantification in sera

Active caspase-1 (Casp1p20) and IL-18 levels were evaluated by ELISA assay (R&D Systems) in the serum from patients with COVID-19 or health donors following manufacturer’s instructions. TNF-α, IL-2, IL-4, IL-6, IL-10, IFN-γ, and IL-17 were quantified in the serum from patients with COVID-19 or health donors using a human CBA cytokine kit (Th1/Th2/Th17 Cytokine Kit, BD Biosciences) following manufacturer’s instructions. IL-1β in the tissue culture supernatants of human monocytes or macrophages cells infected with SARS-CoV-2 was quantified by ELISA (R&D Systems) following manufacturer’s instructions.

### Statistics

Statistical significance for the linear analysis was determined by either two-tailed paired or unpaired Student t-test for data that reached normal distribution and Mann-Whitney was used for not normally distributed data. These statistical procedures and graph plots were performed with GraphPad Prism 8.4.2 software. In addition, longitudinal analyses were implemented to describe variation in IL-18 and Casp1p20 production along time, considering patients’ outcomes and sex as fixed factors. These analyses were performed in R (version 4.0.2) using RStudio (version 1.3.1056), and are detailed as R Markdown object at the supplementary material. The patients’ outcomes were divided into 3 categories (37 patients in total, 16 women and 21 men): death (n = 10 individuals, a total of 25 samples), mild recovery (patients that were hospitalized but did not require mechanical ventilation; n = 17 individuals, a total of 53 samples) and critical recovery (patients that required mechanical ventilation at the UCI and recovered; n = 10 individuals, a total of 27 samples). Activation of IL-18 and Casp1 were evaluated separately using full models that considered the interaction of time (‘Day.sampled’) with patients’ outcomes (‘Outcome’) or sex (‘Sex’) and included individuals (‘Patient.ID’) as a random factor to control for repeated measures and individual effects. Normality and homoscedasticity of the dataset were verified and refuted for time series dataset (see RMarkdown object), and analyses were implemented using the glmmTMB package ^20^ in a generalized mixed model’s (GMM’s) approach with ‘gamma’ distribution and ‘log’ link function. Akaike information criterion for finite samples (AICc) was used to select the best models from the full ones using MuMIn ^21^. Models having AICc values within 2 units of the best-fit model were considered to have substantial support ^22^; adequate residuals distributions were confirmed and representative graphics and tables were constructed using the packages DHARMa ^23^, ggeffects ^24^ and broom.mixed ^25^ - see RMarkdown object to access all parameterization in **Supplemental data**.

### Study approval

The procedures followed in the study were approved by the National Ethics Committee, Brazil (CONEP, CAAE: 30248420.9.0000.5440). Written informed consent was obtained from recruited patients.

## Data Availability

The data that support the findings of this study will be made available upon request or in public repositories.

## Acknowledgments

We are grateful to Maira Nakamura, Bárbara Moreira de Carvalho e Silva and Laura Khouri for technical assistance.

## Author Contributions

T.S.R., and D.S.Z. conceived the study. T.S.R., K.S.G.S. A.Y. I., A.B., S.O., L.A., A.V.G., D.B.P., W.A.A. designed the experiments, defined parameters, collected and processed PBMC and sera samples and analyzed data. R.C., J.E.T., D.C.N., M.H.F.L., C.M.S.S., D.B.C. processed PBMC samples. L.D.C. supervised the collection of PBMC samples from patients. T.S.R., R.B.M., I.A.C., M.C.P. performed the experiments with viral infections. A.T.F., S.S.B., L.S., M.N.B. performed minimally invasive autopsy. K.S.G.S., F.P.V. stained and analyzed autopsy tissues. A.Y.I, F.C.B., T.K. performed bioinformatic analyses, designed and conducted statistical analyses of the data. N.B.A., M.C.G., L.P.B., M.I.F.L., M.N.B., R.C.S., F.C.V., M.A., R.L., S.C.L.A., F.R.O., R.D.R.O., P.L assisted in patient recruitment, collected patient specimens and the epidemiological and clinical data. P.L. supervised and R.D.R.O. helped clinical data management. T. M.C, J.C.A., F.Q.C., L.D.C., F.G.F., T.K., A.T.F., E.A., R.D.R.O., P.L., helped with interpretations of the data. T.S.R., and D.S.Z. drafted the manuscript. All authors helped editing the manuscript. D.S.Z. secured funds and supervised the project.

---

**Fig. S1.**
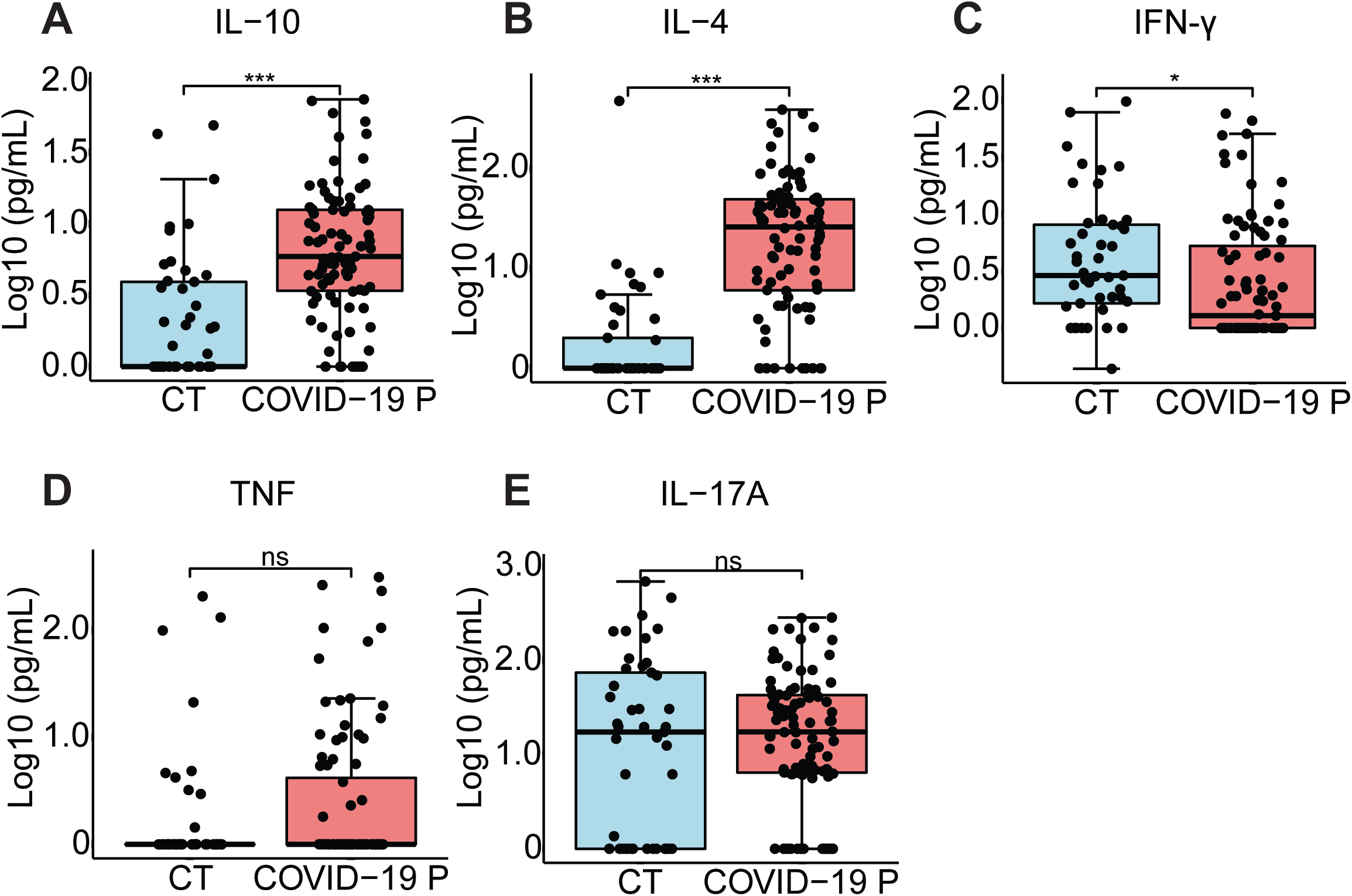
Cytokine production in COVID-19 patients. Cytokine concentration in the serum control individuals (CT, n=45) and COVID-19 patients (COVID-19 P, n=92; all tested positive using RT-PCR). IL-10 (A), IL-4 (B), IFN-γ (C), TNF-α (D) and IL-17A (E) were measured by CBA. Data are shown as Log10-transformed concentrations in pg/mL. ** *P* < 0.01 and *** *P* < 0.001 as determined by Student’s t test. Each dot represents the value form a single individual. Box show average ± SD of the values.

**Fig. S2.**
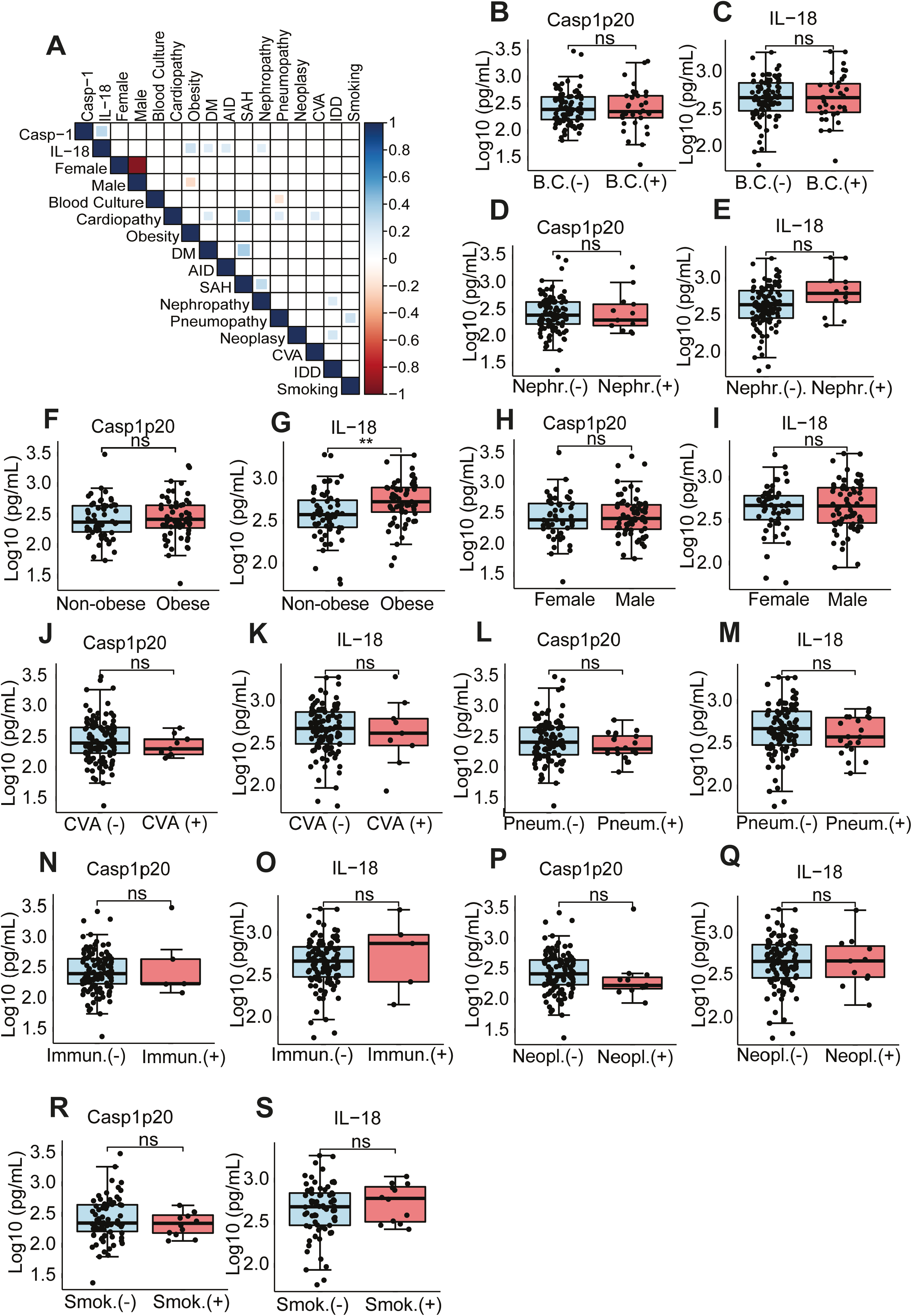
Association of inflammasome activation with clinical characteristics and comorbidities. (A) Matrix correlation of Casp1p20 and IL-18 levels in the serum of COVID-19 patients at the hospitalization day with clinical parameters and comorbidities. (B-S) Levels of Casp1p20 (B, D, F, H, J, L, N, P, R) and IL-18 (C, E, G, I, K, M, O, Q, S) in patients with clinical parameters such as cultivable bacteria in the blood (B, C), nephropathy (D, E), obesity (F, G), gender (H, I), cerebrovascular accident (J, K), pneumopathy (L, M), immunodeficiency (N, O) and neoplasia (P, Q) and smoking (R, S). The levels of Casp1p20 and IL-18 were measured by ELISA and are shown as Log10-transformed concentrations in pg/mL. ** *P* < 0.01 as determined by Student’s t test. Each dot represents value form a single individual. Box show average ± SD of the values.

**Table S1.** Demographic and clinical characteristics of COVID-19 patients.

| Demographics |  | % |
| --- | --- | --- |
| Number | 124 |  |
| Age (years) | 59.25±18.01 |  |
| Female | 50 | 40% |
| Comorbidities |  |  |
| Hypertension | 61 | 49% |
| Obesity | 61 | 49% |
| Diabetes | 46 | 37% |
| History of smoking | 33 | 26% |
| Heart disease | 23 | 18% |
| Lung disease | 20 | 16% |
| Kidney disease | 13 | 10% |
| Cancer | 11 | 8% |
| History of stroke | 9 | 7% |
| Immunodeficiency | 6 | 4% |
| Autoimmune diseases | 2 | 1% |
| Laboratorial findings |  |  |
| CRP (mg/dL)* | 12.55±8.95 |  |
| D-Dimers (µg/mL)** | 2.47±2.59 |  |
| LDH (U/L)# | 565.147±325.9 |  |
| Ferritin (ng/mL)& | 1225.07±1762.9 |  |
| Haemoglobin (g/dL) | 12.31±2.34 |  |
| Neutrophils (cell/mm<sup>3</sup>) | 6728.443±3903.57 |  |
| Lymphocytes (cell/mm<sup>3</sup>) | 1307.37±789.1 |  |
| Platelets (count/mm<sup>3</sup>) | 253426.2±111616.3 |  |
| Medications |  |  |
| Heparin | 112 | 90% |
| Antibiotics | 110 | 88% |
| Glucocorticoids | 58 | 46% |
| Oseltamivir | 56 | 45% |
| Antimalarial | 45 | 36% |
| Respiratory status |  |  |
| Mechanical ventilation | 56 | 45% |
| Nasal-cannula oxygen | 111 | 89% |
| pO<sub>2</sub> | 77.77±29.17 |  |
| SatO<sub>2</sub> | 93.37±5.72 |  |
| Disease Severity |  |  |
| Mild | 7 | 6% |
| Moderate | 49 | 39% |
| Severe | 68 | 55% |
| Outcome |  |  |
| Deaths | 34 | 27% |

\*CRP: C-reactive protein (Normal value <0.5 mg/dL); \*\*D-dimers (NV <0.5 µg/mL); #LDH: lactate dehydrogenase (Normal range: 120-246 U/L); &Ferritin (NR: 10-291 ng/mL)

